# Occupational lead exposure in gasoline station forecourt attendants and other occupations in relation to ALS (amyotrophic lateral sclerosis) risk

**DOI:** 10.1101/2023.05.27.23290632

**Authors:** Lassi Koski, Unathi A. Tshoni, Joshua O. Olowoyo, Aprilia S. Kobyana, Ntebo G. Lion, Liziwe L. Mugivhisa, Sebastian K.T.S. Wärmländer, Per M. Roos

## Abstract

Amyotrophic lateral sclerosis (ALS) is an always fatal neurodegenerative disease characterised by a gradual death of motor neurons in the spinal cord and brain. The cause of ALS is unknown. There appears to be both a genetic and an environmental aspect of ALS disease causation. Multiple occupations are associated with an elevated ALS risk. Interestingly, many of these occupations involve exposure to lead, indicating a possible connection between ALS and lead exposure. Examples include mechanical workers, military service occupations, leather workers and tanners. Gasoline station forecourt attendants, also known as petrol station assistants, show a severely increased ALS risk, and our measurements on forecourt attendants in South Africa show that they display significantly elevated blood lead concentrations. Thus, forecourt attendants can be added to the list of occupations with overlapping risks for lead exposure and ALS incidence. These occupational risks for forecourt attendants are expected to decrease in the future, as leaded gasoline is being phased out worldwide. Nonetheless, the overlapping risks for ALS incidence and lead exposure among forecourt attendants and several other occupations strengthen the hypothesis that lead exposure contributes to ALS.

## 1. Introduction

Amyotrophic lateral sclerosis (ALS) is an always fatal neurodegenerative disease characterised by a gradual death of motor neurons in the spinal cord and brain (Brown and Al-Chalabi, 2017). Worldwide ALS incidence is some 2/100,000 and prevalence 3-5 per 100,000 inhabitants (Brown and Al-Chalabi, 2017), with a significant geographical variation (Masrori and Van Damme, 2020). ALS is characterised by muscle weakness and atrophy leading to respiratory failure and death on average 3 years after disease onset, with a wide variation in survival time (McCombe et al., 2020). A family history of ALS can be found in about 10% of cases (Akcimen et al., 2023). Some 30 various gene-associations have so far been identified within this group (Goutman et al., 2022). Genome-wide association studies have shown a possible genetic component of about 21 % in ALS (Keller et al., 2014). When every case of ALS is considered, the genetic influence on sporadic ALS can be described as weak (Ahmeti et al., 2013). In the light of these conclusions, environmental risk factors for ALS need attention (Ingre et al., 2015).

Various occupational exposures have been explored as preceding events leading to ALS (Gunnarsson and Bodin, 2018). Interestingly, many of these occupations involve exposure to lead (Pb), indicating a possible connection between ALS and Pb exposure. Examples of such occupations with Pb exposure are **military service occupations** (Seals et al., 2016; Peters et al., 2017), **constructions workers** (Fang et al., 2009; Dickerson et al., 2018), **mechanical workers** (Sutedja et al., 2009; Park et al., 2005), **precision tool manufacturers**, and **glass, pottery, and tile workers** (Peters et al., 2017). All these occupations have also been associated with an increased ALS risk (Dickerson et al., 2018; Fang et al., 2009; Park et al., 2005; Peters et al., 2017; Seals et al., 2016; Sutedja et al., 2009). A connection between occupational Pb exposure and elevated ALS risk can furthermore be found for **leather workers** (Buckley et al., 1983) and **tanners** (Bhuiyan et al., 2011; Chio et al., 1991). **Farmers** show an elevated ALS risk (Rosati et al., 1977; Bale, 1975), and farming exposes the farm workers to pesticides and insecticides (Breland and Currier, 1967; Greenberg et al., 1979). Many insecticides have been Pb compounds, especially older preparations in heavy use until the 1960’s, and lead arsenate (PbHAsO_4_) was often prepared by the farm workers themselves by reacting soluble lead salts with sodium arsenate (NaH_2_AsO_4_) (Denver, 2004). Lead arsenate as an insecticide was withdrawn in 1988 (Denver, 2004).

**Gasoline station forecourt attendants**, also known as petrol station assistants, filling station petrol dispensing attendants, fuel pump filling workers, or simply fuel station workers, have previously been shown to have one of the highest occupational ALS risks ever calculated, i.e. OR 8.31; 95% CI 1.79 to 38.54 (Chen et al., 2019). No explanation has so far been provided for this elevated ALS risk among fuel station workers. Here, we report elevated Pb concentrations in blood from forecourt attendants in South Africa, and discuss how this finding strengthens the hypothesis that ALS is induced by Pb exposure.

## 2. ALS in gasoline station forecourt attendants

An extensive population-based study on the association between occupation and ALS risk was conducted in New Zealand between the years 2013 and 2016 (Chen et al., 2019). In a case-control design 321 cases of ALS were compared to 605 controls randomly selected from the Electoral Roll. Occupational history and duration of employment was collected through questionnaires and interviews. An elevated ALS risk was noted for several occupations such as farmers, fishery workers, hunters, trappers, electricians, and telecommunications technicians (Chen et al., 2019). The common denominator for these varied occupations remains to be elucidated. The highest odds ratio (OR) for ALS risk was calculated for the job title classification 52113 i.e. forecourt attendants (OR 8.31, 95% CI 1.79 to 38.54). This OR is 2-3 times higher than any other occupation with elevated ALS risk in this large study, and the upper limit of the confidence interval reaches OR 38 even though this OR is based on only 11 cases (Chen et al., 2019). This number should be seen in perspective of the low ALS prevalence in New Zealand, i.e. 2.3/100000 (Cao et al., 2018). Prior to the New Zealand study on Motor Neurone Disease Association (Chen et al., 2019), petrol station forecourt attendants were not included with their own code in the New Zealand classification of occupations. Before this re-classification, the specific exposure to Pb from gasoline might have been spread out over other occupational classes, making an ALS risk OR of 8.31 for forecourt attendants even more exceptional. A slightly lower risk, but still high, was documented for the job title G5321 i.e. automotive fuel retailing (Chen et al., 2019). It can therefore be suspected that exposure to automotive fuel, e.g. gasoline/petrol, is responsible for the increased ALS risks among both gasoline station forecourt attendants and automotive fuel retailers.

## 3. Materials and Methods

### 3.1 Participants

The study participants (n=74) lived in Gauteng, a suburb to Pretoria in South Africa. The study involved 38 individuals who were either currently or previously employed as gasoline station forecourt attendants. This group was divided into subgroups based on years of service, as shown in Table 1. As controls served 36 individuals who did not have any contact with petrol on a daily basis, i.e. students or workers who lived far away from any of the studied petrol stations, and who reached their workplace on foot. A written consent form was signed by all participants, who also filled out a structured questionnaire, with details on occupation, health status, smoking habits, and sociodemographic status. Ethical approval for the project was provided by the Sefako Makgatho Health Sciences Research Committee (SMUREC/S/91/2020:PG).

**Table 1.** Lead concentrations [μg/L] in blood from forecourt attendants in relation to number of years worked. Reprinted from reference: (Olowoyo et al., 2023).

|  | Minimum | Maximum | Mean ± SD |
| --- | --- | --- | --- |
| 1-3 years of service | 5.21 | 28.5 | 10.5 ± 5.9 |
| 4-6 years of service | 7.41 | 23.8 | 14.3 ± 4.9 |
| 7 and more years of service | 5.04 | 60.2 | 15.1 ± 12.9 |
| Control group | 4.37 | 41.3 | 16.9 ± 9.2 |

### 3.2 Gasoline station forecourt attendant job description

In South Africa, the daily duties of the forecourt attendants are important to the management of the gasoline station, as they represent the station business establishment and as such have certain obligations. This job is about to disappear as self-service is spreading at gas stations over the world, however in South Africa traditionally the attendant provides manual service by walking out to each vehicle and fuelling it. No protective devices are used between the car gasoline tank and the open air during filling, and the attendants are therefore frequently inhaling gasoline vapour. Along with dispensing gasoline, the employees frequently check the tire pressures and the amount of oil and water in the vehicles. Some gasoline stations in South Africa are open for 24 hours, and work shifts are split between employees to meet all hours. A typical shift for a gasoline station forecourt attendant lasts about 8 hours, but supervisory staff members may work up to 12 hours.

### 3.3 Analysis of metal concentrations

Methods and steps for collecting the blood samples and analysing their concentrations of Pb have been described previously (Olowoyo et al., 2023). In short, blood was collected from an antecubital vein of each participant by qualified health workers from Dr. George Mukhari Academic Hospital Pretoria, using established procedures. The blood samples were digested by 0.5% HCl and 1.5% HN0_3_ in purified water. The digested sample was then diluted with an internal standard solution, centrifuged, and Pb concentrations in the supernatants measured with an Agilent 7700 series inductively coupled plasma mass spectrometry instrument (Olowoyo et al., 2021). The blood sample analysis was conducted by an accredited laboratory in South Africa, where the samples were analysed in triplicate using ClinChek® certified reference materials. Statistical analysis was performed with the SPSS 28.0 statistical software. One-way ANOVA tests were used to determine if the differences in Pb concentrations between different study groups were statistically significant or not. The level of statistical significance was set to p<0.05.

## 4. Results

Blood Pb concentrations of petrol station forecourt attendants are significantly elevated compared to controls. Lead concentrations, years of exposure, and lifestyle habits associated with the work situation, are presented in Tables 1-4 below.

The overall Pb concentrations in blood of these forecourt attendants have previously been reported (Olowoyo et al., 2023), and those results are here reprinted in Table 1. Blood Pb concentrations increase with number of years worked, and the difference between the group with 7 or more years of service (Pb = 15.1 μg/L) and the group with 1-3 years of service (Pb = 10.5 μg/L) is statistically significant (p<0.05).

Table 2 shows that the blood Pb concentrations are significantly higher (p<0.05) in forecourt attendants that are smokers (Pb = 19.5 μg/L) compared to those that are non-smokers (Pb = 10.2 μg/L). This is not unexpected, as it is well known that tobacco products contain toxic metals including lead (Bernhard et al., 2005; Pappas et al., 2016; Wallin et al., 2017).

**Table 2.** Lead concentrations [μg/L] in blood from forecourt attendants who are smokers and non-smokers, respectively.

|  | Minimum | Maximum | Mean ± SD |
| --- | --- | --- | --- |
| Non-smokers | 5.04 | 15.8 | 10.2 ± 3.8 |
| Smokers | 5.04 | 60.2 | 19.5 ± 13.9 |

The lifestyle habits of the forecourt attendants are shown in Table 3. Most of the participants (92%) changed their uniforms after working one or two shifts, but 8% did not. It can be assumed that some petrol products are spilled on the uniform during a normal workday, and re-using a dirty uniform will therefore increase the dermal exposure to petroleum products. Similarly, the 11% of the forecourt attendants that do not shower after work will likely suffer more dermal exposure to petroleum than the 89% that do shower (Table 3). Around half of the participants (45%) lived in the same area where they worked, and these are likely the same 45% of participants who walked to their jobs (Table 3). The remaining 55% of participants commuted to their workplaces via public transportation (52%) or with their own car (3%), thereby exposing themselves to bus and car exhaust fumes, which are known to contain harmful chemicals. For example, elevated Pb concentrations have been reported in the blood of bus drivers in Indonesia (Kurniasih et al., 2018).

**Table 3.** Lifestyle habits associated with the work situation of forecourt attendants.

| Lifestyle habits |  | N=38 |
| --- | --- | --- |
| Uniform change | 1 wear | 16 (42%) |
|  | 2 wears | 19 (50%) |
|  | 3 wears | 2 (5%) |
|  | After a week | 1 (3%) |
| Shower after work | Yes | 34 (89%) |
|  | No | 4 (11%) |
| Live in the area | Yes | 17 (45%) |
|  | No | 21 (55%) |
| Transportation | Own car | 1 (3%) |
|  | Public transport | 20 (52%) |
|  | Walking | 17 (45%) |

## 5. Discussion

The causes and origins of ALS have been debated for more than a century, and still no clear conclusions can be drawn (Ingre et al., 2015). In an attempt to correlate various occupational exposures to ALS risk, 321 ALS cases in New Zealand were studied with a full occupational history and compared to 605 controls (Chen et al., 2019). Very high elevated risks were observed for gasoline station forecourt attendants and automotive fuel retailers, suggesting that exposure to automotive fuel, e.g. gasoline/petrol, is responsible for these increased ALS risks. This possibility is supported by another more recent New Zealand study showing that occupational exposure to gasoline was associated with an elevated (OR=2.24) ALS risk (Chen et al., 2022). Few studies have however investigated gasoline as a possible trigger for ALS.

Gasoline/petrol is a liquid mainly consisting of hydrocarbons in the form of alkanes, alkenes, and aromatic components many of which are neurotoxic and/or carcinogenic (Eisler, 2000; Sholts et al., 2017). There are also some reports that organic substances, including aromatic ones, may contribute to ALS (Malek et al., 2015). The possible mechanisms are unclear, but one study has found that organic molecules such as aromatics and dioxins that bind to the aryl hydrocarbon receptor (AHR), increase production of TDP-43 (Ash et al., 2017), a protein that appears to be involved in ALS pathology (Mackenzie et al., 2007; Berning and Walker, 2019; Koski et al., 2021). But gasoline contains also other substances, such as trace levels of metals including Fe, Zn, and Pb (Kitto, 1993; Chrastny et al., 2018).

During the 20^th^ c., external Pb was often added to gasoline to improve the performance of combustion engines (Angrand et al., 2022). The use of Pb additives has gradually been phased out, and as a result blood Pb concentrations have decreased worldwide (Angrand et al., 2022). In South Africa, Pb additives were banned from automobile gasoline in 2006 (Olowoyo et al., 2023), and in 2021 Algeria became the last country on the planet to ban Pb additives in automobile gasoline (Angrand et al., 2022). However, Pb is still a common additive in gasoline for aeroplanes, off-road motorbikes, racing cars, and military vehicles. The intrinsic Pb in non-leaded gasoline, which typically can be differentiated from added Pb due to different isotope compositions, should furthermore not be ignored as it can contribute non-negligible amounts of Pb to the environment (Chrastny et al., 2018).

Our measurements on forecourt attendants in South Africa (Olowoyo et al., 2023) show that these workers display significantly elevated blood Pb concentrations (Table 1). The underlying Pb exposure is most certainly work-related, as the Pb concentrations increase with work years (Table 1). However, other factors such as cigarette smoking also contribute to Pb exposure (Table 2), and work-related life-style habits (Table 3) do influence the amount of Pb exposure at work. These results are not surprising, as earlier studies have found significantly elevated blood Pb concentrations in fuel station workers in Iraq (Al-Rudainy, 2010), India (Bawaskar and Bawaskar, 2020), Nigeria (Alli, 2015), Sudan (Tayrab et al., 2014) and Iran (Bahrami et al., 2002).

As Pb is an established risk factor for ALS, the higher blood Pb levels observed in forecourt attendants, suggest that Pb exposure is at least a partial explanation as to why forecourt assistants and other occupations that handle gasoline are more likely to develop ALS. This is consistent with the observation that some of the earliest described ALS cases were related to Pb exposure (Roos, 2013; Aran, 1850; Darwall, 1831). In fact, long-time low dose Pb exposure produces a clinical picture indistinguishable from ALS (Campbell et al., 1970; Livesley and Sissons, 1968). In Australia, some 250,000 tonnes of Pb were emitted from gasoline between 1933 and 2002, (i.e. until leaded automobile gasoline was banned), and an Australian study found that lifetime Pb exposure from gasoline was associated with age-specific ALS death rates (Zahran et al., 2017). A related study used linear regression models to estimate a temporal association between accumulated gasoline lead emissions and ALS in Australia. That study found a best fit correlation between a 20-year lag of gasoline lead emissions and percent all cause ALS death (R2 = 0.98, p = 2.6 × 10^−44^) (Laidlaw et al., 2015). In the coming decades, we expect to see reduced risks for Pb exposure as well as ALS incidence in forecourt attendants, due to both the removal of lead additives in automobile gasoline and the increasing number of self-service fuel stations across the world.

The forecourt attendants in the current study, however, were exposed to petrol fumes containing Pb in their daily working environment. Lead in petrol fumes is inhaled into the lung alveoli and to some extent deposited in the lungs, however the majority of Pb passes the alveolar endothelium and capillary endothelium into the bloodstream. This respiratory exposure is the main Pb exposure route for forecourt attendants, but Pb can also pass the skin barrier and enter the bloodstream directly after dermal exposure (Nordberg and Costa, 2021). Lead is transported in the blood tightly bound to haemoglobin in red blood cells (Simons, 1986) and distributed to every organ system, mostly kidneys, liver and bone (Nordberg and Costa, 2021). Lead is to 90 % stored in bone in adults (Flora et al., 2012) and from bone it can re-enter into the circulation in response to changes in bone mineralization (Silbergeld et al., 1993). Blood Pb reaches the cerebral arteries, arterioles and capillaries, where it can traverse the capillary endothelia which constitute the blood-brain barrier into the cerebral interstitial fluid (Lossinsky and Shivers, 2004). Alternatively, it can directly pass the blood-CSF barrier at the level of the choroid plexus into the cerebrospinal fluid (CSF) which surrounds the cerebral cortex and the spinal cord. Lead concentrations in CSF are normally low, but Pb can also accumulate in the peripheral nervous system in exposed individuals (Nordberg and Costa, 2021). Lead is classified as a selective choroid plexus toxicant, and Pb accumulates to a great extent in the choroid plexus (Manton and Cook, 1984; Zheng, 2001) and disturbs its barrier functions (Zheng, 2001). Spinal cord anterior horn motor neurons, the nerve cells primarily affected in ALS, seem to be specifically vulnerable to Pb toxicity (Limonta and Capellini, 1976; da Silva et al., 2020; Beritic, 1989; Roos, 2013) and to selenite toxicity (Raber et al., 2010; Vinceti et al., 2012). The details regarding how Pb might trigger ALS are unclear, and several mechanisms are possible. For example, Pb ions do induce oxidative stress, mimic other metal ions, and modulate the aggregation of harmful amyloid proteins such as amyloid-β (Wallin et al., 2017) and possibly also TDP-43 (Koski et al., 2021). In evaluating the direct toxic effects of Pb on the anterior horn cells of the spinal cord in ALS patients, the actual blood Pb concentration itself might be less important than the concentration ratios between Pb and other neurotoxic and/or protective metals (Koski et al., 2023) that can contribute to synergistic or attenuating effects in relation to Pb.

## 6. Conclusions

Gasoline station forecourt attendants display significantly elevated blood Pb concentrations, both due to respiratory Pb exposure from gasoline and due to lifestyle habits, such as cigarette smoking. The overlapping risks of ALS incidence and Pb exposure among forecourt attendants might be causally connected, in the sense that occupational Pb exposure arguably is one of the factors that contribute to ALS in these workers. The same could be true for many of the other occupations where Pb exposure and increased ALS risk coexist.

## Data Availability

All data produced in the present work are contained in the manuscript.

## Acknowledgments

Support to SW from the Magnus Bergvall foundation and support to PMR from the Kamprad Research Foundation, the Magnus Bergvall Research Foundation, the Ulla-Carin Lindquist Foundation for ALS Research and the Karolinska Institutet IMM strategic grants is gratefully acknowledged. The support from NRF, South Africa is also greatly appreciated.

## Conflicts of interest

The authors declare no conflict of interest.

## Notes

### Competing Interest Statement

The authors have declared no competing interest.

### Author Declarations

Ethical approval for the project was provided by the Sefako Makgatho Health Sciences Research Committee (SMUREC/S/91/2020:PG)

